# Rising prevalence of *Plasmodium falciparum* artemisinin resistance mutations in Ethiopia

**DOI:** 10.1101/2024.09.11.24313421

**Authors:** Bokretsion G. Brhane, Abebe A. Fola, Helen Nigussie, Alec Leonetti, Moges Kassa, Henok Hailgiorgis, Yonas Wuletaw, Adugna Abera, Hussein Mohammed, Heven Sime, Abeba G/Tsadik, Gudissa Assefa, Hiwot Solomon, Geremew Tasew, Getachew Tollera, Mesay Hailu, Jonathan J. Juliano, Ashenafi Assefa, Jonathan B. Parr, Jeffrey A. Bailey

## Abstract

Ethiopia is striving to eliminate local malaria transmission by 2030, despite a recent resurgence of malaria cases due to multiple factors. A significant contributor to this resurgence could be drug resistance, particularly the emergence of partial resistance to artemisinin (ArtR) in Ethiopia and other regions of Eastern Africa. This situation highlights the necessity for genomic surveillance to monitor relevant drug resistance markers. This study reports sentinel site-based genomic surveillance results for *P. falciparum* antimalarial drug resistance mutations. From 2019 to 2022, dried blood spots (DBS) were collected from febrile outpatients ≥1 year of age with microscopically confirmed falciparum malaria at 12 sentinel sites across 5 regions. Molecular inversion probe (MIP) sequencing targeted mutations associated with artemisinin and partner drug resistance, including *k13*, *mdr1, crt, dhfr,* and *dhps* genes, along with genome-wide markers to assess the complexity of infection (COI) and parasite relatedness. A total of 1,199 falciparum*-*positive patients were assessed, with a median age of 20 years (IQR: 14-30) and including 463 (38.6%) females. The WHO-validated K13 R622I mutation had a high but regionally variable prevalence (15.7%, range 0-58.8%). The validated K13 A675V mutation was detected for the first time in Ethiopia in the Gambella Region (4.5%), as well as P441L and P574L mutations were detected at low frequencies in Southern and Oromia Regions, respectively. Several partner drug resistance markers were identified, with mutations in MDR1(184F), DHPS, DHFR, and CRT nearly fixed across the country. Most samples (87.2%) were monogenic infections (COI=1) and showed high genetic relatedness, particularly within the health facilities. Principal component analysis revealed regional clustering of parasites, particularly in Gambella. The prevalence of K13 R622I across the country and the presence of multiple additional ArtR markers emphasizes the urgent need for rigorous monitoring of artemisinin combination therapy (ACT) efficacy to detect partner drug resistance and ACT failure early and its impact on malaria resurgence in Ethiopia.

## Background

Malaria remains a significant public health challenge in Africa, accounting for approximately 95% of malaria cases and 96% of malaria deaths, predominantly affecting children under five and pregnant women^1^. Despite notable progress through the use of artemisinin-based combination therapies (ACTs), vector control measures, and vaccines like RTS,S, the path to eliminating malaria faces formidable obstacles ^2,3^. These challenges include the emergence of drug- and diagnostic-resistant parasites^4–7^, insecticide-resistant mosquitoes^8^, new invasive mosquito species^9^, inadequate healthcare infrastructure, and funding gaps^10^. Resistance to artemisinin poses a particular threat to the efficacy of ACTs in Africa^11,12^ and progress towards malaria elimination, highlighting the need for new strategies to define and mitigate the impact of antimalarial resistance.

Artemisinin partial resistance (ArtR) was first reported in western Cambodia in Southeast Asia in 2006-2007^13,14^ and has since spread or emerged independently in other countries in the Greater Mekong Subregion^15^, South America^16^, and recently in Eastern Africa^17–19^ and the Horn of Africa^5,20^. The rise of ArtR WHO-validated markers have been confirmed in Ethiopia^5^, Eritrea^20^, Rwanda^17^, Uganda^21^, Tanzania^6^, and Democratic Republic of the Congo (DRC)^22^. ArtR likely accelerates the emergence of resistance to partner drugs^23^, and once combined with partner drug resistance lead to significant clinical failure in Southeast Asia. Frank partner drug resistance mutations have not been found in Africa. Amino acid polymorphisms in the multidrug resistance protein 1 (MDR1) and chloroquine resistance transporter (CRT) are well established and partially effect susceptibility diametrically to both amodiaquine and possibly lumefantrine^24^, which are the most common partner drugs used in ACTs in Ethiopia and most of Africa. This decreased susceptibility likely further undermines ACTs increasing the potential for emergence of other mutations^4,6^.

Despite Ethiopia’s gains in malaria control over the past two decades, the country faces increasing malaria cases in various regions, in the setting multiple challenges: First, the K13 622I mutation, a validated marker of partial artemisinin resistance, has been found at significant frequencies, particularly in the north of the country^5^. Second, the rise of *P. falciparum* strains with *hrp2* and *hrp3* gene deletions complicates the reliability of rapid diagnostic tests (RDTs), contributing to diagnostic failures and potentially increasing transmission^5,7^. Third, invasive urban-adapted mosquito species^9,25^, and increasing insecticide resistance threaten malaria control efforts^26^. Fourth, environmental and man-made factors such as climate change^27^, conflict, and population movement may have facilitated the spread of malaria, contributing to the recent resurgence. Gaps in surveillance systems hinder effective tracking of disease patterns and resistance trends and understanding the role of these challenges in malaria resurgence. Dissecting and addressing these challenges requires enhanced surveillance, robust diagnostic tools, and innovative strategies to track and mitigate the spread of drug-resistant strains to achieve elimination goals^28^. Genomic surveillance of molecular markers of resistance and understanding the genomic diversity of the parasite will be critical^29^. Such surveillance helps identify resistance hotspots and informs targeted interventions such as deployment of sequential or triple ACT regimens, contributing to more effective malaria control and elimination strategies^30,31^.

Integrating genomic data with routine surveillance systems for timely tracking of emerging and spreading resistance is crucial^32,33^. To this end, the Ethiopian Public Health Institute (EPHI), in collaboration with the national malaria control and elimination program, established a network of sentinel sites representative of the diverse malaria transmission ecologies across the country. These sites collect and analyze data on malaria transmission dynamics, treatment efficacy, and resistance patterns, providing opportunities for evidence-based decision-making in near real-time. These sentinel sites are integral to Ethiopia’s broader malaria control and elimination strategy, which aims to reduce the disease’s burden through early detection, targeted treatment, and prevention efforts. This study reports an in-depth analysis of the emergence and spread of drug resistance mutations and parasite relatedness using molecular inversion probe (MIP) sequencing of samples collected from 12 sentinel sites across the country.

## Results

### Study population

After filtering, 855 of 1,199 samples were included in the analysis sample set (**Figure S1**). Samples came from participants enrolled at all 12 sentinel site health facilities (**Figure 1A**) across five regions (Amhara = 270, Gambella = 131, Oromia = 193, SNNP = 221, Somali = 40) (**Figure 1B**). The median parasite density of sequenced samples was 2,000 parasites/µL (range 48 to 61,000 parasites/µL) by microscopy, with better sequencing coverage in samples with higher parasitemia (**Figure S1**). Most samples (n = 767) were collected in 2022 and 79.2% (n = 607) were successfully genotyped, 393 collected in 2019 and 56.9% (n=224) successfully genotyped followed 39 collected in 2023 and 61.5% (n = 24) successfully genotyped. Details of samples successfully sequenced per each MIP panel summarized in **Table S1** and **Table S2.**

**Figure 1.**
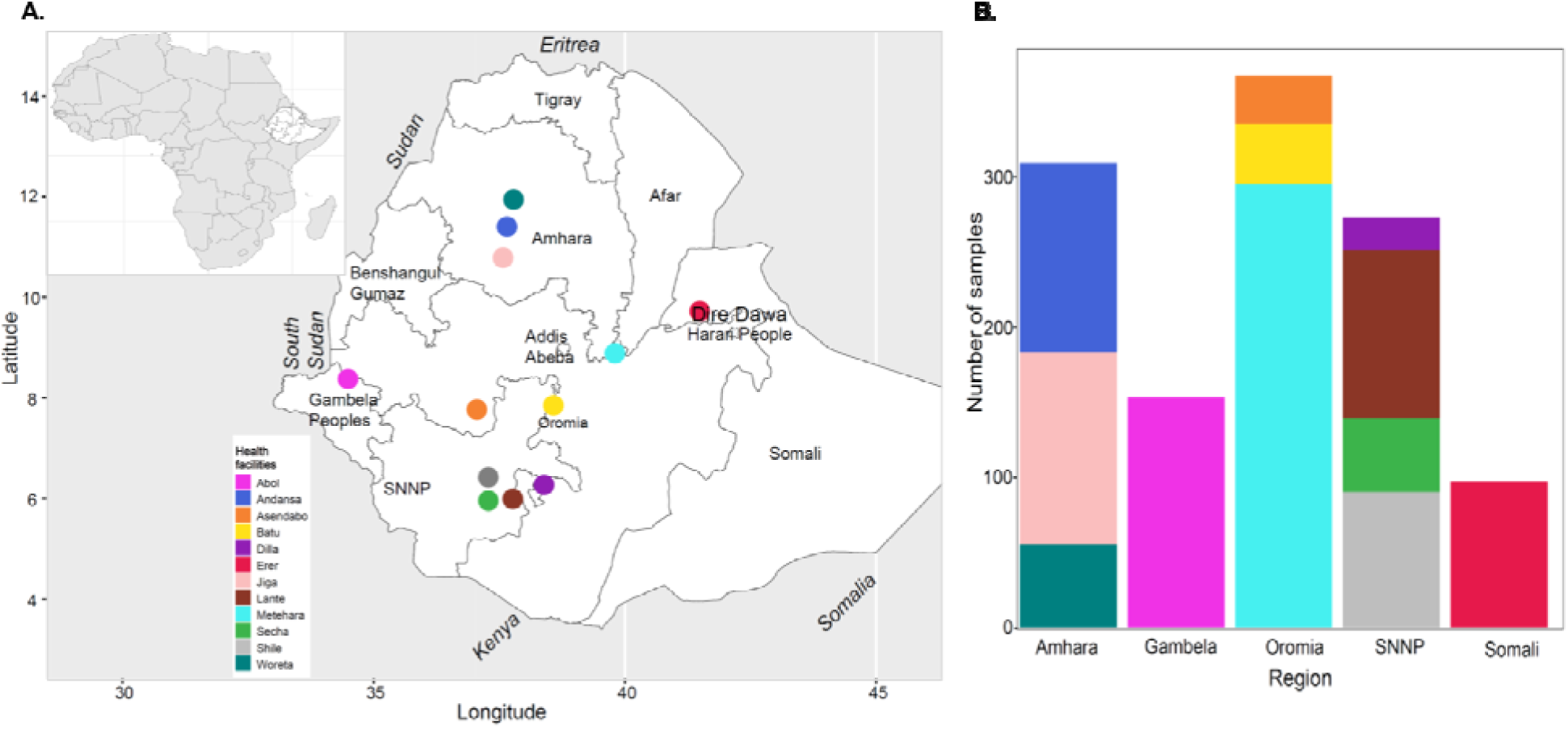
Study sites and sequenced samples. **A)** Study sites and **B)** distribution of samples sequenced per health facility and region. Colors indicate individual health facilities.

### Prevalence and distribution of ArtR mutations

We first assessed the prevalence of WHO-validated and candidate K13 ArtR mutations revealing four WHO validated K13 mutations (**Figure 2A**). K13 R622I was the most prevalent (15.7% [95% CI 13.2-18.2%]). Its geospatial distribution was heterogeneous (prevalence range: 0 to 58.8%) at health facility level (**Figure S2**), with highest regional prevalence in the north (46.9% [95% CI 40.7-53.2%] in the Amhara region) where it was first reported at 2.4% in one facility in 2014^34^. K13 R622I was also prevalent in Erer (12.9%), a Somali region Eastern part of the country, and additionally observed in low frequency from health facilities from Gambela and Oromia regions but not reported from health facilities from SNNP (Southern Nations, Nationalities and Peoples) region.

**Figure 2.**
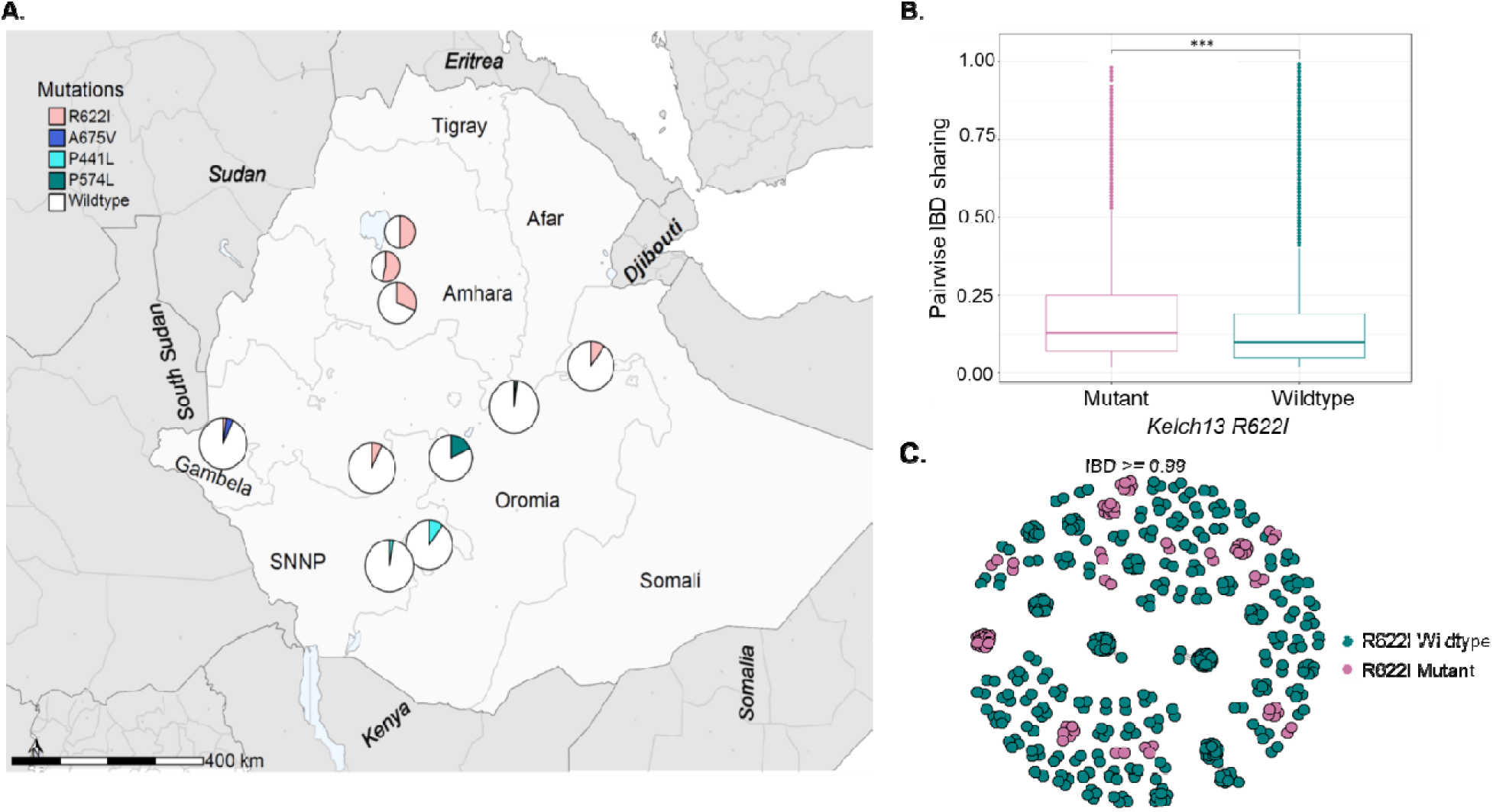
WHO-validated K13 mutation prevalences and relatedness of 622I mutant vs wildtype parasites. **A)** Spatial distribution of WHO-validated K13 mutations by health facility (pie charts). Colors indicate which mutation, and pie chart size is proportional to sample size per study site. **B)** Pairwise IBD sharing within *kelch13* R622I mutant versus wild parasites, showing *** significant differences (*P* < 0.001, two-tailed, Mann–Whitney *U*-test). Boxes indicate the interquartile range, the line indicates the median, the whiskers show the 95% confidence intervals and dots show outlier values. *P* value determined using Mann–Whitney test is shown. **C)** Network analysis showing highly related parasite pairs (IBD≥0.99). Each node identifies a unique isolate, and an edge is drawn between two isolates if they meet the IBD threshold. Isolates that do not have a IBD pairwise connection above threshold are not shown. Color codes correspond to K13 R622I mutant (pink) and wild parasites (blue green).

Of concern, we found three additional WHO-validated K13 mutations not previously observed in the Horn of Africa or Ethiopia in the southern half of the country where 622I is not as prevalent. We found 6 samples with A675V (Gambella Region), 4 samples with P441L (3 from Southern Region including 2 from Dilla and 1 from Secha health centers, and 1 from Oromia Region in the Metehara health center) and 5 samples with P574L (Oromia Region in the Batu health center) (**Table S3**). We also identified an additional 14 non-synonymous mutations within and outside the K13 propeller domain with variable frequencies (**Figure S3**). Other K13 mutations not associated with ArtR were found, including K189T at 32% (CI, 27.1-38.3) prevalence across sites.

### Genetic relatedness of k13 mutant parasites

To further elucidate the spatial spread of mutant parasites, we conducted IBD relatedness analysis and found that there was a higher average IBD sharing estimate for K13 R622I mutant isolates compared to K13 wild-type isolates (**Figure 2B**). This was also true for parasites carrying P574L mutations (**Figure S4B**), however, parasites carrying A675V or P441L did not show significant relatedness and were in fact less related compared to wildtype (**Figure S4A, C**). We then conducted relatedness network analysis, using an IBD cut of ≥0.99 to detect evidence of clonal transmission, finding clustering mutant parasites clustered into pairs and networks (**Figure 2C**) separate from wildtype.

### Prevalence of mutations associated resistance to partner drugs

Mutations in the *P. falciparum* multidrug-resistance gene 1 (MDR1), particularly isolates that carry the N86 (wild), 184F (mutant), and D1246 (wild) haplotype have been associated with modest decreased susceptibility to lumefantrine (the first line partner drug in Ethiopia, co-formulated as artemether-lumefantrine). Our analysis revealed that 99.1% ([95% CI 98.4-99.6%]) sequenced samples carried N86 (wild), followed by 98.1% ([95% CI 97.2-98.9%]) samples of D1246 (wild), and 94.8% ([95% CI 93.4-96.3%]) carrying 184F (mutant) (**Figure 3A**). The overall high prevalence resulted in less spatial variation at district or health center level (data not shown). Nearly all parasites with the ArtR K13 R622I (**Figure 3A**) and A567V (**Figure 3B**) mutations also carry the MDR1 NFD haplotype associated with lumefantrine resistance. The co-occurrence of ArtR and partner drug resistance mutations raises concern about the effectiveness of ACTs in Ethiopia.

**Figure 3.**
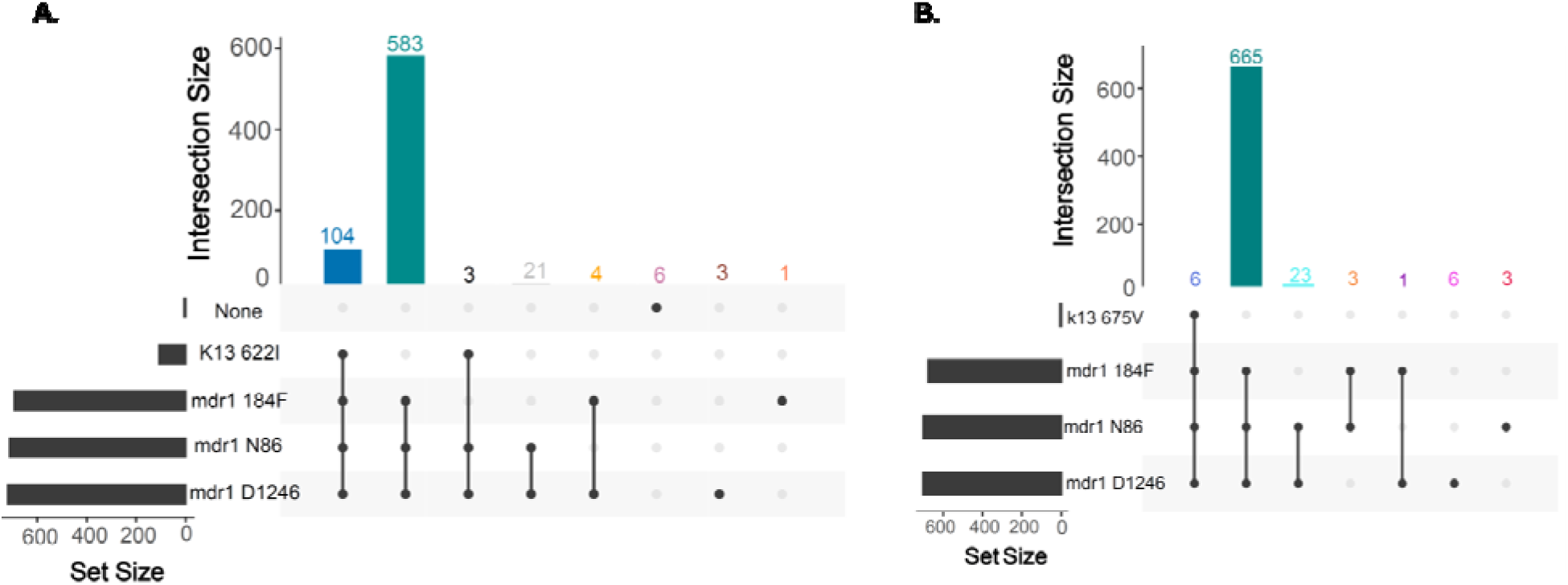
Co-occurrence of ArtR and lumefantrine-resistance markers in Ethiopia. Number of parasites with **A)** K13 622I and MDR1 N86 (wild), 184F (mutant) and D1246 (wild) and with **B)** K13 675V and MDR1 N86 (wild), 184F (mutant) and D1246 (wild). Only samples with calls across all loci are shown. For polygenomic infections, the dominant haplotype (≥51% allele frequency) is shown.

We also observed a high prevalence of mutations associated with resistance to sulfadoxine-pyrimethamine (SP) and chloroquine. SP was discontinued in 2004 and is not used for intermittent preventive treatment for pregnant women (IPTp) as in other African countries. Nonetheless, SP resistance mutations persist at high prevalence. In DHFR, 98.9% ([95% CI 98.6-99.4%]) parasites had the S108N mutation, 98.5% ([95% CI 97.8-99.3%]) N51I, and 65% ([95% CI 61.9-68.1%]) C59R (**Figure S5A**) mutations associated with pyrimethamine resistance. Overall, 469 (61.6%) samples carry all three mutations (DHFR IRN, triple mutant) with low spatial heterogeneity at regional level across the country. None of the genotyped samples carried DHFR 164L, a mutation associated with higher levels of SP resistance. Analysis of markers associated with sulfadoxine resistance revealed that all 907 (100%) isolates carried DHPS A437G and 83.1% ([95% CI 80.6-85.4%]) K540E, with 711 (78.8%) carrying both of these mutations (DHPS GE, double mutant) (**Figure S5A**). The DHPS A581G mutation, shown to increase SP resistance, when co-occurring with the above mutations occurred at low frequency, 3.4% ([95% CI 2.6-5.4%]) (**Figure S5A**).

Chloroquine is widely used in Ethiopia for treatment of *Plasmodium vivax* infection. Mutations in the *P. falciparum* chloroquine (CQ) resistance transporter (CRT) were common; 896 (82.4%) parasites had the chloroquine-resistance marker 76T. All regions had high prevalence with the exception of Gambela in the southwest 7.6% (([95% CI 5.2-8.4%]) (**Figure S5B**). Additional mutations in CRT are provided in **Table S3**.

### Complexity of infection and parasite population structure

The majority of sequenced samples were monogenomic 745 (87.1%) (**Figure S6A**), consistent with previous reports^5,35^, with some level variation in complexity of infection at the regional level (**Figure 4A**). The maximum COI detected was 4 (from Gambella Region) likely reflecting relatively low transmission in the study areas. Principal components (PC) 1 and 2 revealed distinct clustering of parasites from Gambella, supporting regional differentiation of the *P. falciparum* population within Ethiopia, but not by K13 622I status (**Figure 4B**, **Figure S7**).

**Figure 4.**
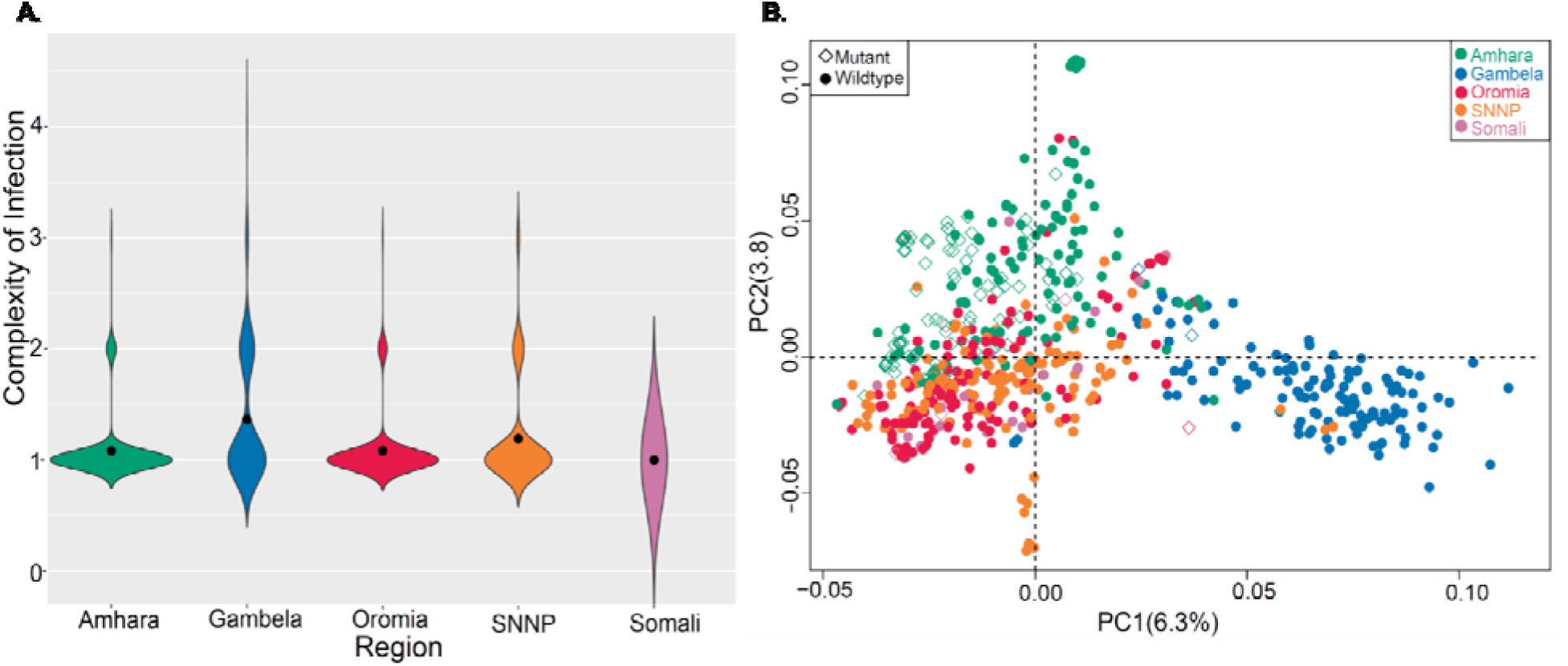
Complexity of infection and population structure of *P. falciparum* in Ethiopia. **A**) Complexity of infections for *P. falciparum* at a regional level. Colors indicate regions and black dots inside violin plots indicate median COI. **B**) PCA plot of *P. falciparum* population at regional level. Colors indicate geography of sample origins and shapes indicate mutation K13 R622I status. Each diamond or dot indicates individual parasites. Percentage of variance explained by each PCA presented in each plot.

### Spatial patterns of parasite relatedness and connectivity

We conducted IBD analysis of monogenomic infections to further elucidate the connectivity of the parasite population across different regions. Overall, we observed a tailed distribution of highly related parasite pairs (**Figure 5A**) with some level of spatial heterogeneity of proportion of pairs IBD for parasite population at each health center (**Figure 5B**). The lowest average IBD sharing observed at Abol health center, Gambella Region. We then conducted relatedness networks analysis to assess where haplotypes sharing IBD above the specified threshold cluster geographic region or sample origin and found clear clustering of highly related parasite pairs (IBD≥0.90) within health centers suggesting inbreeding and clonal transmission at local scale (**Figure 5C**). However, occasional sharing of nearly clonal parasites was seen between clinics, suggesting long range parasite sharing. To identify if transmission of highly related parasite pairs (IBD≥0.90) were occurring between clinics in a within a region and between different regions, we visualized a relatedness network on top of the Ethiopia map and found that majority of the highly related pairs were distributed within regions or nearby regions than distant regions. Connections between Gambella and other regions were rare (**Figure 5D**).

**Figure 5.**
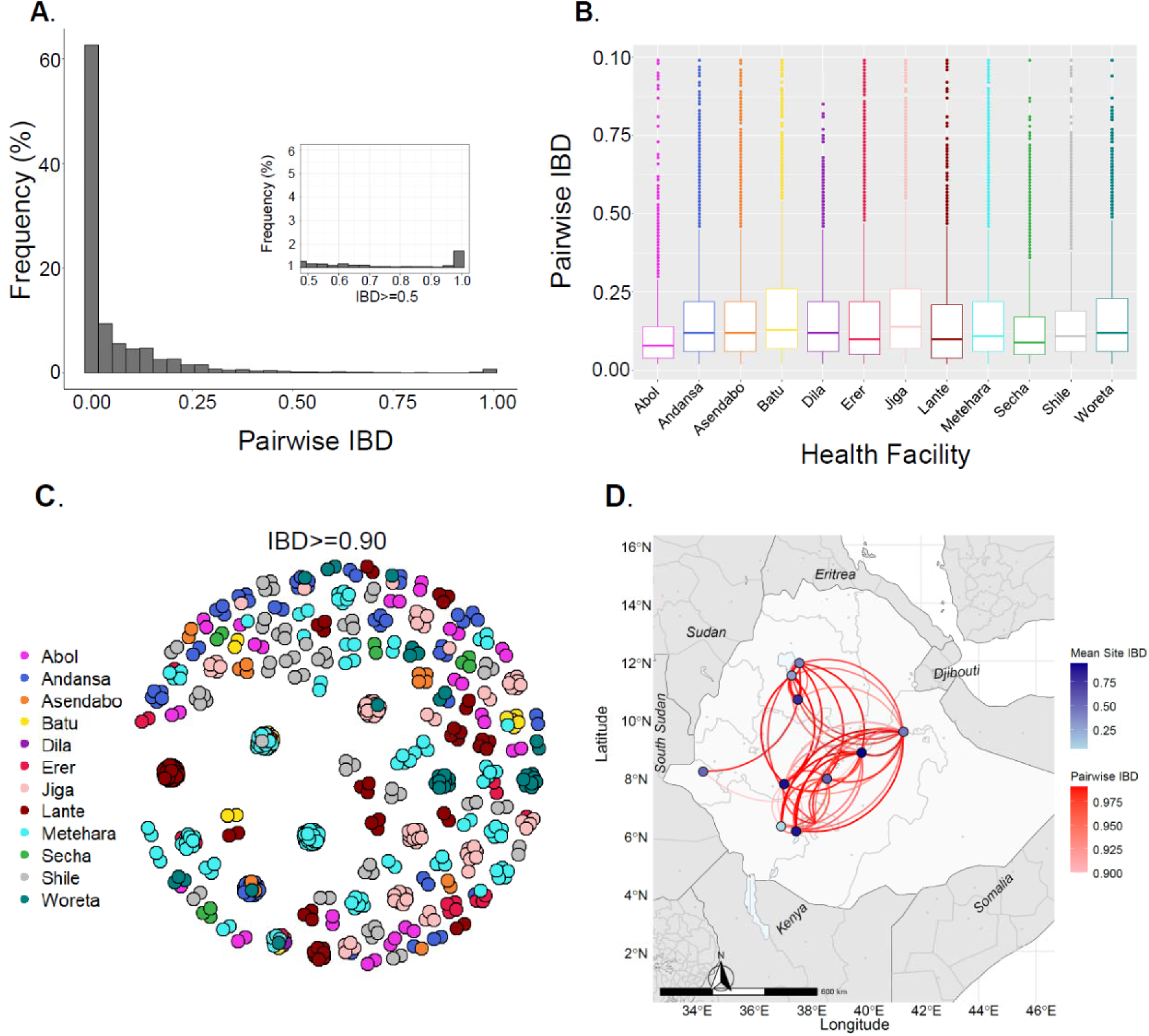
*P. falciparum* parasite relatedness and connectivity across Ethiopia. **A)** Pairwise IBD sharing across 12 health facilities. The plot shows the probability that any two isolates are identical by descent, where the *x* axis indicates IBD values and the *y* axis indicates the frequency (%) of isolates sharing IBD. The inset highlights highly related parasite pairs with a heavy tail in the distribution and some highly related pairs of samples having IBD ≥ 0.90. **B)** Pairwise IBD sharing within a health facility. Boxes indicate the interquartile range, the line indicates the median, the whiskers show the 95% confidence intervals and dots show outlier values. **C)** Relatedness network of highly related parasite pairs (IBD ≥ 0.90) at study site. Each dot indicates a sample and colors correspond to health facilities across five regions in Ethiopia. Each node identifies a unique isolate and an edge is drawn between two isolates if they share their genome above IBD ≥ 0.90. Isolates that do not share IBD ≥ 0.90 of their genomes with any other isolates are not shown. **D)** Spatial distribution of highly related parasite pairs with IBD ≥ 0.90. Color of circles or connecting lines represents the level of IBD sharing within and between sites, respectively. The within site shading is based upon the most related pair of parasites at the site.

## Discussion

Findings from our genomic analysis of parasites collected across sentinel sites predominantly in 2022 suggest a critical threat of ArtR to ACT effectiveness in Ethiopia. We identified increased prevalence and evidence of spread of the previously identified WHO-validated ArtR K13 622I, with marked increases compared to previous reports. Here we see a prevalence of 622I up to 53.7% in Andansa Health Centre within the Amhara region, as compared to 10% in 2018^5^ and 2.4% in 2014^34^. Parasites carrying 622I are more closely related and clustered at health facility/district level compared to wildtype parasites.

These findings are consistent with our previous report of clonal transmission in three regions of Ethiopia^5^. The highly-related, clonal nature of K13 622I mutant parasites compared to wildtype parasites in this study is consistent with inbreeding and increasing frequency within populations, especially in low transmission areas^36,37^. Our findings highlight the value of establishing genomic surveillance at malaria sentinel sites, particularly how it can play a crucial role in tracking the emergence and spread of drug-resistant strains by providing continuous and localized monitoring of malaria parasite populations. It also provides deeper insight into the genetic population structure of parasites circulating at these sites. Researchers in collaboration with the malaria control program can detect early signals of resistance development, understand its geographical spread, and assess the effectiveness of current treatments^38,39^.

In addition to K13 622I, three WHO-validated K13 mutations (A675V from Gambella Region; P441L from South and Oromia Regions, and P574L from Oromia Region) are reported for the first time in Ethiopia. These K13 mutations are becoming more common in East Africa^18,40–42^. The K13 441L mutation found in three health centers from different geographic areas with variable malaria transmission and a recent increase in malaria cases. The validated K13 P574L mutation was present in 5 samples from only one site, Batu health center in the Oromia region. K13 P574L mutations have each been shown to have multiple independent origins throughout SE Asia^43^ and have been reported in Africa^44^.

The K13 675V mutation was only found in one health center from Gambella Region of western Ethiopia neighboring South Sudan. This mutation was reported in isolates collected in Uganda beginning in 2016. It has proven to enhance parasite survival *in vitro* and has been associated with resistance *in vitro*^42,45^. Importation of the 675V mutation seems probable given the IBD analysis showed lower relatedness and 4/6 of the participants with the mutation reported travel. However, we cannot confirm whether the haplotype carrying this mutation are imported or locally emerged due lack of publicly available WGS data with K13 675V mutations from East Africa. Potentially supporting the impact of travel and importation on parasites in Ethiopia, the parasite population from Gambella did not cluster with other parasite populations in the country. This differentiation could be for multiple reasons, such as different travel patterns in the country, potentially supported by the low number of long-distance connections with other clinics (**Figure 5D**). However, Gambella also shares a border with South Sudan and receives a large number of refugees from across the border. Recent studies of South Sudanese refugees entering and arriving at the Ugandan Adjumani refugee camp show 15.4% prevalence of 675V^46^. Combined, this suggests that 675V may be spreading rapidly across South Sudan thereby entering Ethiopia.

Different studies propose that high efficacy of partner drugs, high prevalence of polygenomic infections, and high parasite genetic diversity impede the spread of ACT resistance in Africa^21,47,16^. However, in this study we found a high prevalence of mutations associated with lumefantrine resistance^48^. The majority of genotyped samples carried the MDR1 NFD haplotype^49,50^, but three parasites with WHO-validated ArtR markers in our study carry MDR1 NFD haplotypes that could lead to ACT failure. Recent study showed that there is increasing prevalence of MDR1 NFD haplotypes in recent years in East Africa^51^. Moreover, the rollout of ACTs has led to an increase in the MDR1 NFD haplotypes across several African studies, suggesting parasite populations in Africa are under significant ACT selective pressure^49^. With reports of decreased ACT efficacy and treatment failure in Africa^52–54^, the presence of partner drug mutations and high prevalence of monogenomic infections suggests the conditions are ripe for ACT failure in Ethiopia. Thus, close monitoring of ACT efficacy and preparation for deployment of alternative treatment strategies (e.g. multiple sequential therapies, sequential ACT or triple ACT) are needed.

This study does have limitations. First it does not address whether the mutations are directly emerging locally or are imported. This will be possible as more genomic information is produced from other areas of Africa and whole genome sequencing of these isolates is done allowing for accurate tracking of K13 haplotypes. Second, it did not address the impact of these mutations on clinical resistance as the in vitro efficacy was not evaluated. Third, 12 sentinel sites do not represent the entirety of malaria epidemiology or parasite populations circulating in Ethiopia. Nonetheless, sampling across most regions of the country provides valuable, generalizable information for the malaria control program. Despite these limitations, these data provide important findings about how ArtR is spreading in the Horn of Africa which is critical information for malaria control programs. Lastly, our data suggests importation and population movement may play a role in the genetic patterns seen, but we do not have details on locations and duration of travel to test this.

In conclusion, this study highlights critical challenges for malaria control including: 1) the continued emergence and spread of the validated 622I mutation, 2) the detection of new validated ArtR mutations in the Horn of Africa, and 3) the high rate of the co-occurrence of ArtR with the MDR1 NFD haplotype associated with reduced sensitivity to lumefantrine, the key partner drug in the region. It also raises additional questions about the impact of importation and travel on the spread of antimalarial resistance, questions that will be answered as more genomic data from Africa becomes available. We also demonstrate that sentinel surveillance systems can leverage genomics to provide critical information to control programs and potentially can help target where *in vivo* studies should be focused to determine the clinical impacts of antimalarial resistance polymorphisms, as was done for validated ArtR in Tanzania due to the 561H mutation^11,41^. All of East Africa and the Horn of Africa are going to face significant challenges in the years to come due to emerging antimalarial resistance, and malaria molecular surveillance will play a key role in fighting this emerging threat^4,6,55^.

## Methods

### Study sites and sample collection

EPHI maintains 25 malaria sentinel sites, representing the variable eco-epidemiological zones across the country. A series of cross-sectional studies were conducted in 12 of these sites between 2019 and 2022 to assess the emergence and prevalence of antimalarial drug resistance markers. The study sentinel sites included at least one area where large scale industry and irrigation, typically for sugar, is occurring and one location in a border area (**Figure 1A**). From September to November of each year, patients attending the outpatient department with signs and symptoms of malaria were screened by microscopy. Microscopy occurred in the field and was reviewed by a second expert microscopist at EPHI. In cases of discordance, a third reader was used, blinded to previous reads. Inclusion criteria included *P. falciparum* positive by microscopy. Patients less than 1 year or with severe malaria were excluded. Following consent, a brief questionnaire was performed (including patient age, sex, malaria sign and symptoms, travel history and demography), and dried blood spot (DBS) samples were blotted onto Whatman 3 filter paper, air-dried, placed individually in a plastic bag containing silica gel desiccant. These were stored at room temperature prior to transport to EPHI, and stored at −20 °C until used.

### DNA extraction and MIP sequencing

DNA was extracted from DBS (one full DBS spot, typically involving 2 or 3 6mm punches) using Chelex-Tween as previously described^56^.

Five microliters of extracted DNA were used for each of the MIP captures using two panels: one covering key *P. falciparum* drug resistance genes and mutations, including *k13*, *mdr1, crt, dhfr,* and *dhps* genes, and a newly designed panel of 305 MIPs targeting common SNPs (>5%) SNPs **(Table S4)** (Niare *et al.* in preparation). MIP capture and library preparation were performed as previously described^5,57^. Sequencing was conducted using an Illumina NextSeq 550 instrument (150 bp paired-end reads) at Brown University (RI, USA). For samples with newly detected K13 441L, 574L and 675V, mutations in Ethiopia), an additional MIP capture was done using the same MIP panel and resequenced to a high depth to confirm the mutations. Controls for each MIP capture and sequencing included genomic DNA from serial dilution of lab strain 3D7 as well as no template and no probe controls.

### Variant calling and filtering

The sequencing reads generated for each MIP panel were demultiplexed separately using MIPtools software (https://github.com/bailey-lab/MIPTools) and further processed using MIP Wrangler software (https://github.com/bailey-lab/MIPWrangler), in which sequence reads sharing the same Unique Molecular Identifiers (UMIs) were collapsed to generate a single consensus. Each dataset was analyzed by mapping sequence reads to the *P. falciparum* 3D7 reference genome using Burrows-Wheeler Aligner (BWA)^58^. Then variant calling was performed using freebayes software^59^. To reduce false positives due to PCR and alignment errors, the alternative allele needed to be supported by more than one UMI within a sample, and the allele must have been represented by at least 10 UMIs across the entire population. For genome wide SNPs, only biallelic variant SNP positions were retained for downstream analysis. Low quality SNPs (mapping quality < 30), were removed from the analysis using vcftools^60^; additionally, individual variant calls within each sample were set to missing if the site was not supported by at least five UMIs. After these steps, SNPs with more than 50% missing data across samples, and subsequently samples with more than 50% of SNPs missing, were removed from all downstream analyses^5^.

### Drug resistance prevalence estimation

For drug resistance analysis, variants were annotated using the *Pf 3D*7 v3 reference genome and gene features. Variants with a unique UMI count of 3 or greater used to estimate prevalence key and known drug resistance mutations associated with different antimalarial resistance using the *miplicorn* package R 4.2.1 software (https://github.com/bailey-lab/miplicorn) and custom R script. With the exception of the DHPS 437 mutation (which is mutant in the reference genome), drug resistance prevalence was determined considering both heterozygous or homozygous variants as mutant alleles, while homozygous reference alleles considered as wild-type, and missing loci not included for prevalence calculation (p=x/n*100), where p = prevalence, x = number of mutant alleles, n = number of successfully genotyped loci. n is different for each locus as each locus had independent success rates for genotyping. Prevalence and frequency bar plots were generated using the *ggplot2* package and spatial visualization of prevalence on maps of study areas were created using the *sf* package in R 4.2.1 software. Analysis of haplotypes included only samples where complete genotypes across all involved variant sites were available and was plotted and visualized using the *UpSet* package in R 4.2.1 software. Finally, the prevalence of each drug resistance marker was calculated at the health facility level.

### Analysis of complexity of infectious and parasite population structure

The complexity of infection (COI) is the number of distinct parasite clones infecting a single host. To estimate the number of clones per sample, the VCF file containing SNP data was converted to THE REAL McCOIL categorical method format: heterozygous call as 0.5, homozygous reference allele as 0, homozygous alternative allele as 1 and no call as −1 and used as an input file for analysis of COI using THE REAL McCOIL R package. To assess whether parasite populations within Ethiopia clustered per their geographic origin, or by mutation status, we conducted principal component analysis (PCA) using *SNPRelate* function in R 4.2.1 software and the result was visualized ggplot2.

### Analysis of parasite relatedness using Identity-by-descent (IBD)

We used identity-by-descent (IBD) to measure relatedness between parasites and identify regions of the genome shared with recent common ancestry using the *inbreeding_mle* function in MIPAnalyser software (v.1.0.1) (https://github.com/mrc-ide/MIPanalyzer) in R 4.2.1 software as previously described ^61^. In brief, both monogenomic and polygenomic infections (major alleles used for heterozygous positions) were included in the IBD analysis. Inbreeding_mle uses a Markov chain Monte Carlo to detect genomic regions that are identical by descent (IBD) and aids simultaneous detection of parasite population clustering. Networks of highly-related parasites per k13 mutation status or geographic origin (health facilities) created using the *igraph* package in R 4.2.1 software. We then assessed IBD sharing at the regional and local (health facility level) scales in order to assess spatial distribution of the parasite population.

## Data availability

All data produced in the present study are available upon reasonable request to the authors.

## Ethics

Before the commencement of the study, scientific and ethical approval was obtained from the Ethiopian Public Health Institute’s Institutional Review Board (EPHI-IRB), protocol number EPHI-IRB-398-2021. In addition, respective permission and supporting letters were obtained from the health facilities administration. All the necessary precautionary and ethical methods were considered during the implementation of the study for the patients and investigators. All screening forms and case record forms are kept in a secured location, with access limited to authorized staff members. Unique numerical identifiers were used for the computer-based data entry and blood samples. Genotyping work conducted at Brown University was considered non-human subjects research. Only aggregated clinical data and de-identified samples used for reporting individual level genotypes for publication.

## Author contributions

BGB, AA, AAF, JBP, JJJ and JAB conceived the study. BGB led patient recruitment and sample collection with contributions from HN, MK, HH, YW, GT, AA, MH, HS, AG, GA, HS, GT and AA. BGB, AL and AAF performed laboratory work. BGB and AAF led genetic data analysis and wrote the first draft of the manuscript. AA, JBP, JJJ and JAB supported genetic data analysis and interpretations of results. All authors contributed to the writing of the manuscript and approved the final version before submission.

## Data Availability

All data produced in the present study are available upon reasonable request to the authors.

## Acknowledgments

The authors used an artificial intelligence language model for English language editing during the writing process. However, the manuscript is original to the authors, who take responsibility for its content.

## Supplementary materials

### Supplementary Figures

**Figure S1.**
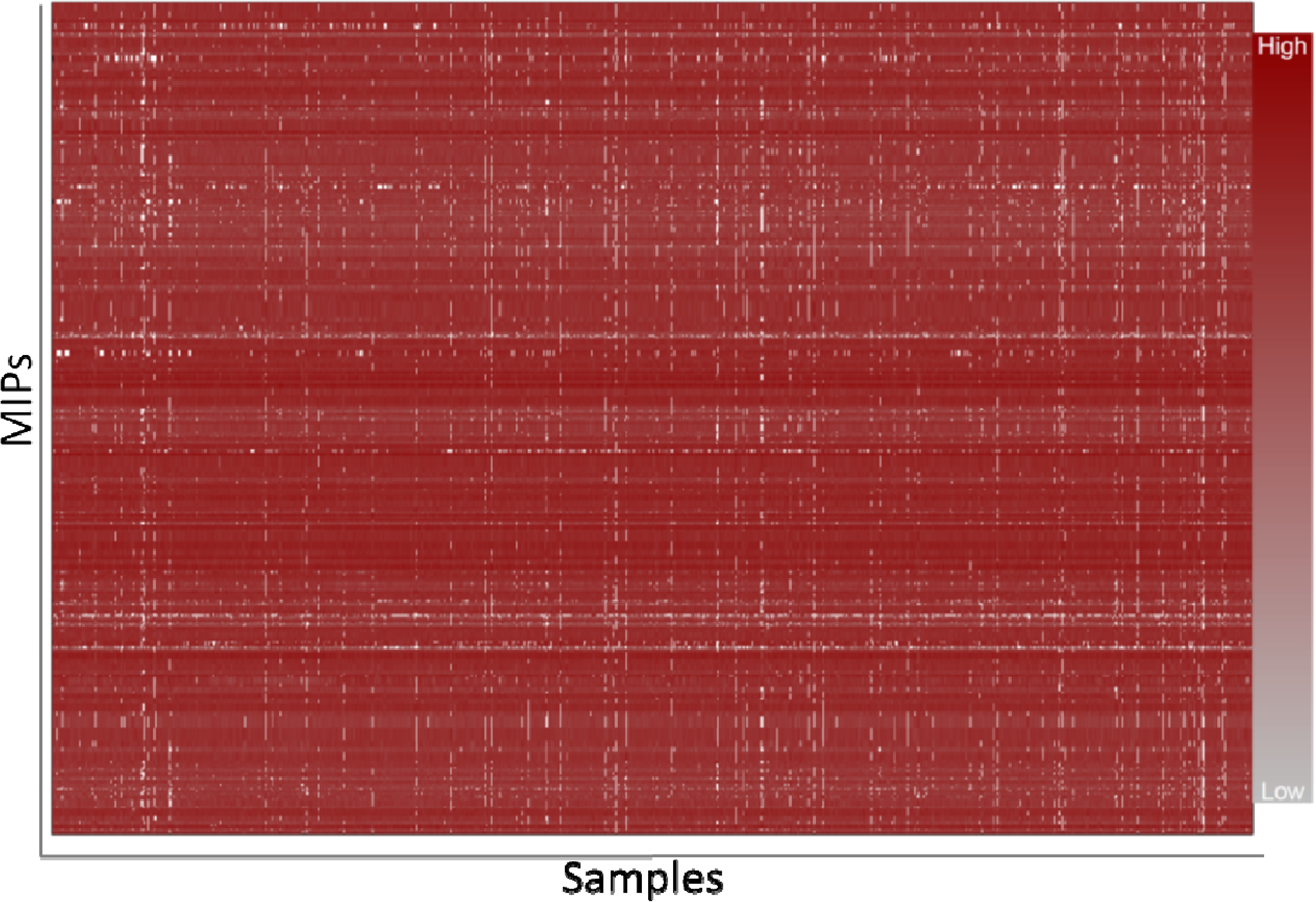
MIP sequencing coverage for genome-wide SNPs. Heatmap color shows samples (n = 855, columns) and loci (n = 1415, rows) coverage retained after filtering for downstream analysis.

**Figure S2.**
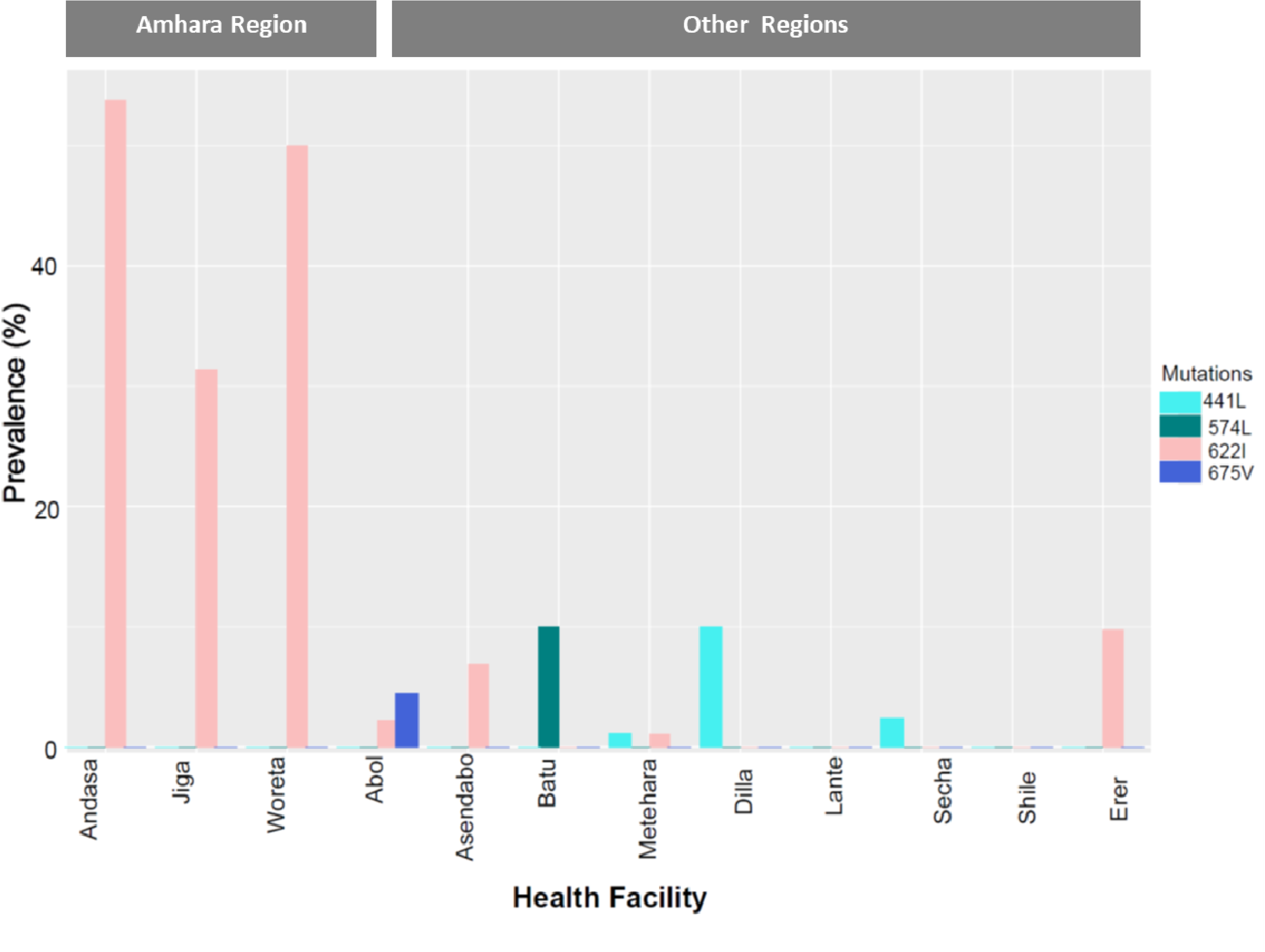
WHO validated K13 mutations prevalence at health facility level. Bar plots show prevalence of the indicated mutations at health facility level per region. Colours indicate mutations

**Figure S3.**
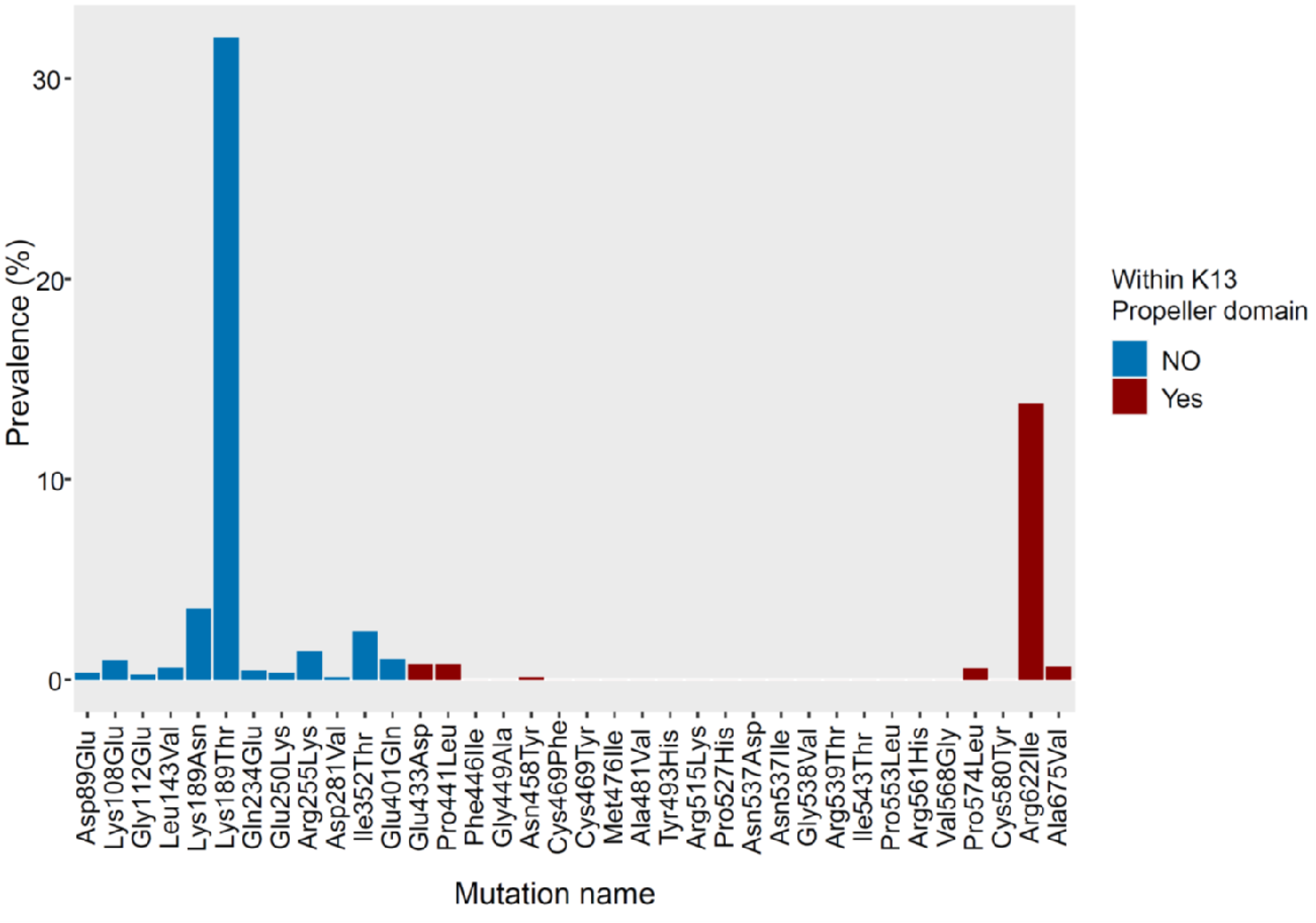
Prevalence of non-synonymous mutations across the *K13* gene, coloured according to amino-acid residues within beta propeller domain where validated resistance mutations are located or not.

**Figure S4.**
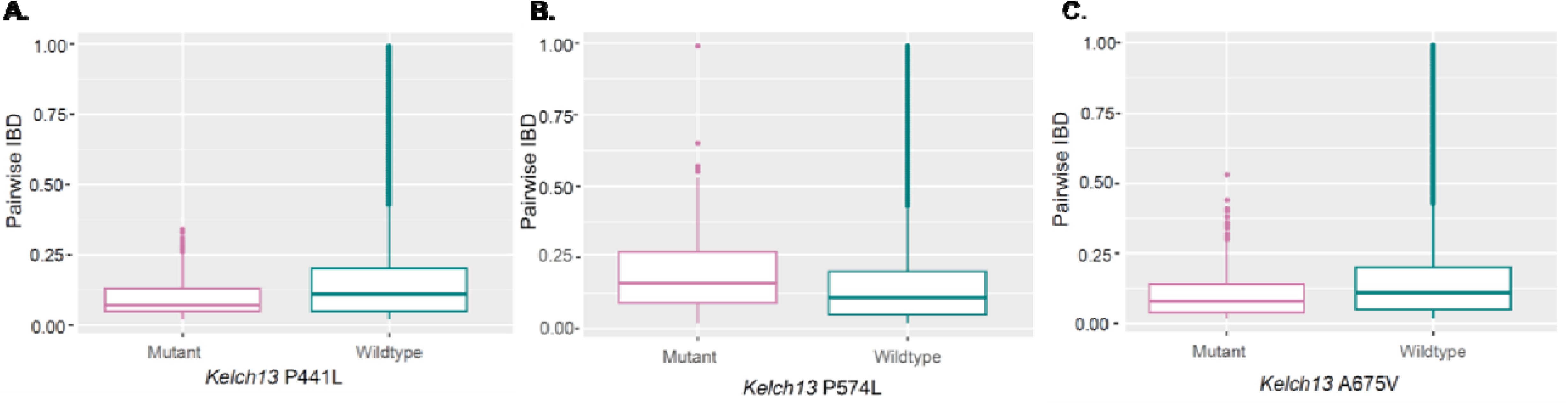
Comparison of relatedness of mutant vs wildtype. **A)** Plot shows the mean IBD within K13 P441L mutant versus wildtype parasites. **B**) Plot shows the mean IBD within K13 P574L mutant versus wild type parasites. **C**) Plot shows the mean IBD within K13 A675V mutant versus wild type parasites. Color codes correspond to *kelch13* R622I mutant and wild parasites.

**Fig S5.**
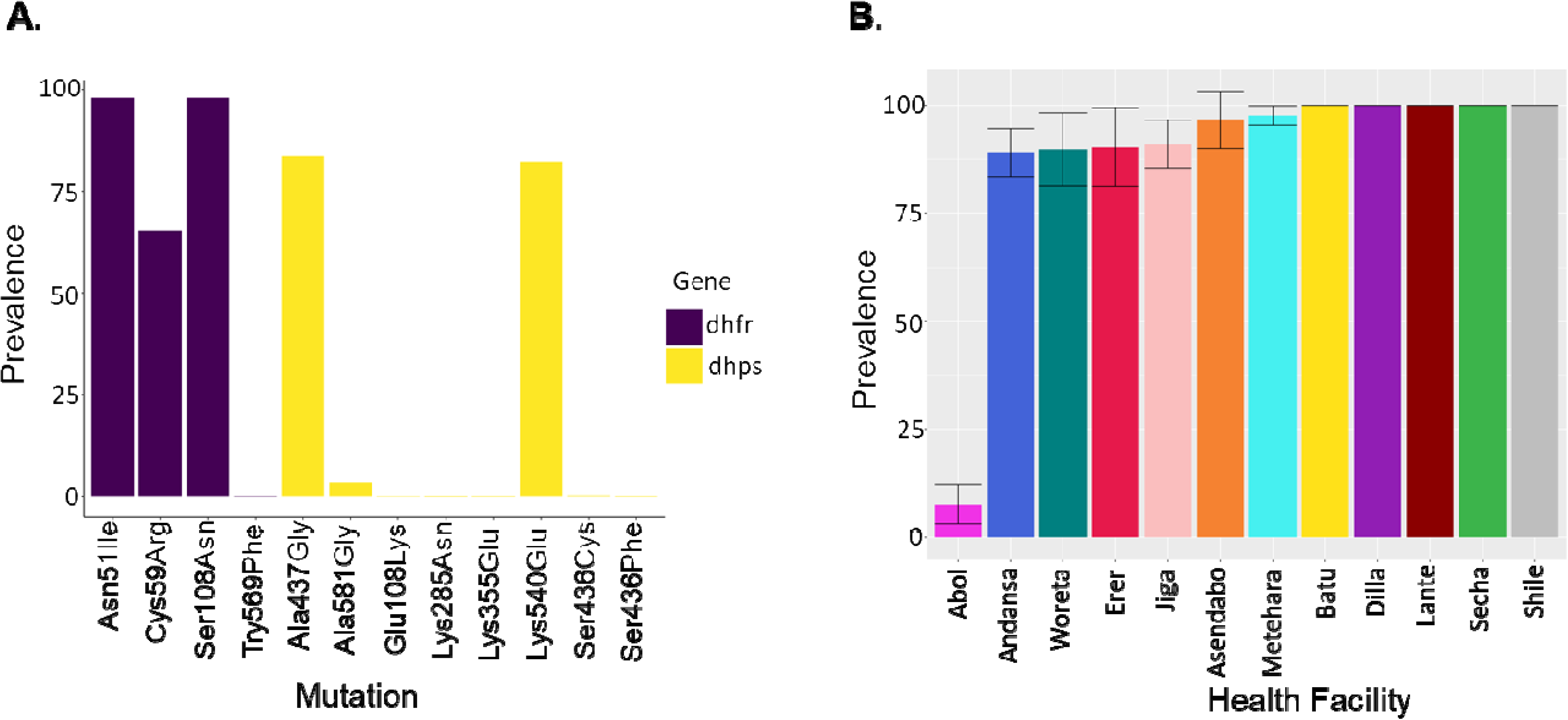
Prevalence of SP (A) and CRT K76T (B) mutations in Ethiopia. **A)** commutative prevalence of different SP markers. Bars indicate individual mutation and colors indicate gene. **B)** Prevalence of CRT K76T prevalence at health facility level.

**Figure S6.**
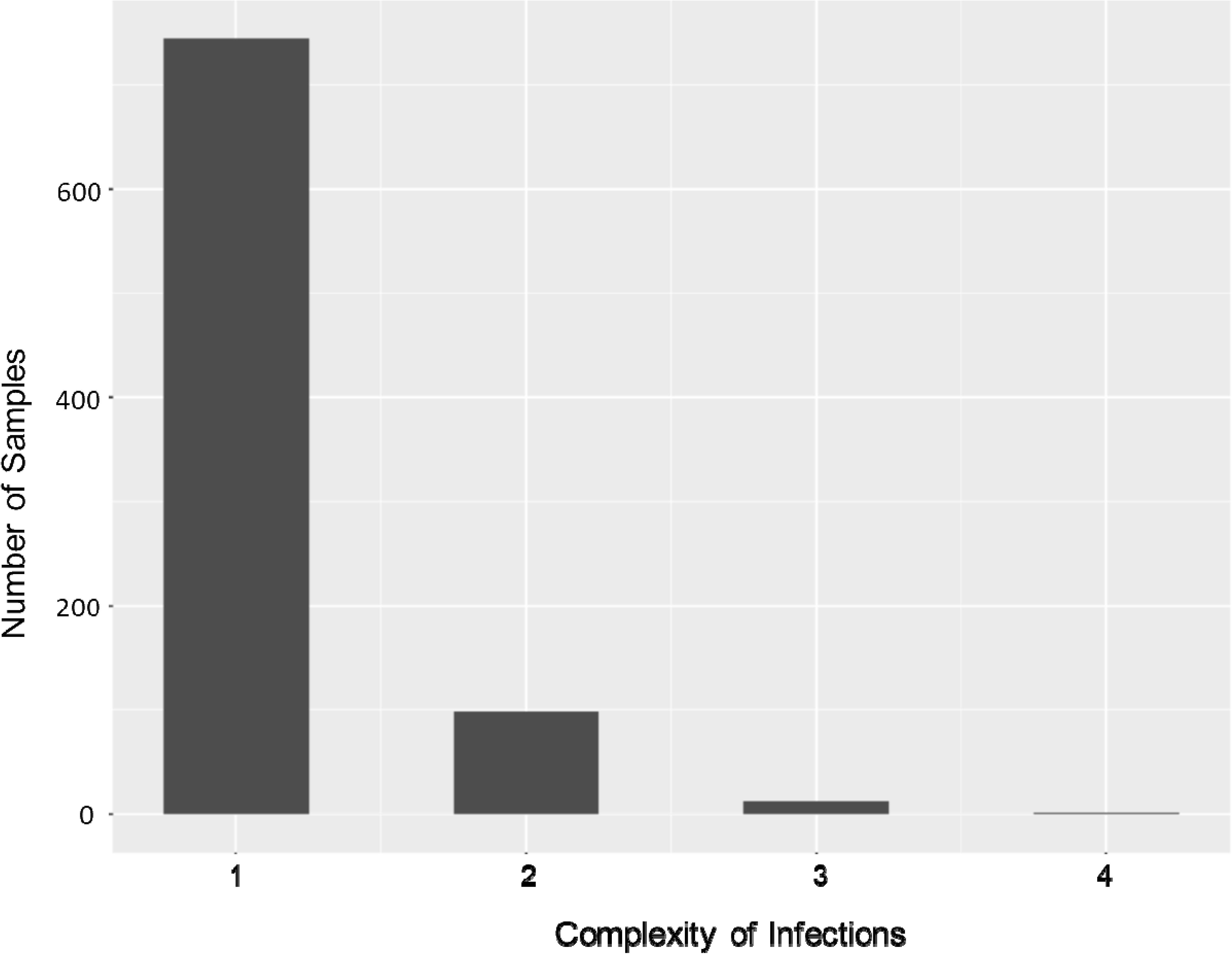
Complexity of infections across sequenced samples.

**Figure S7.**
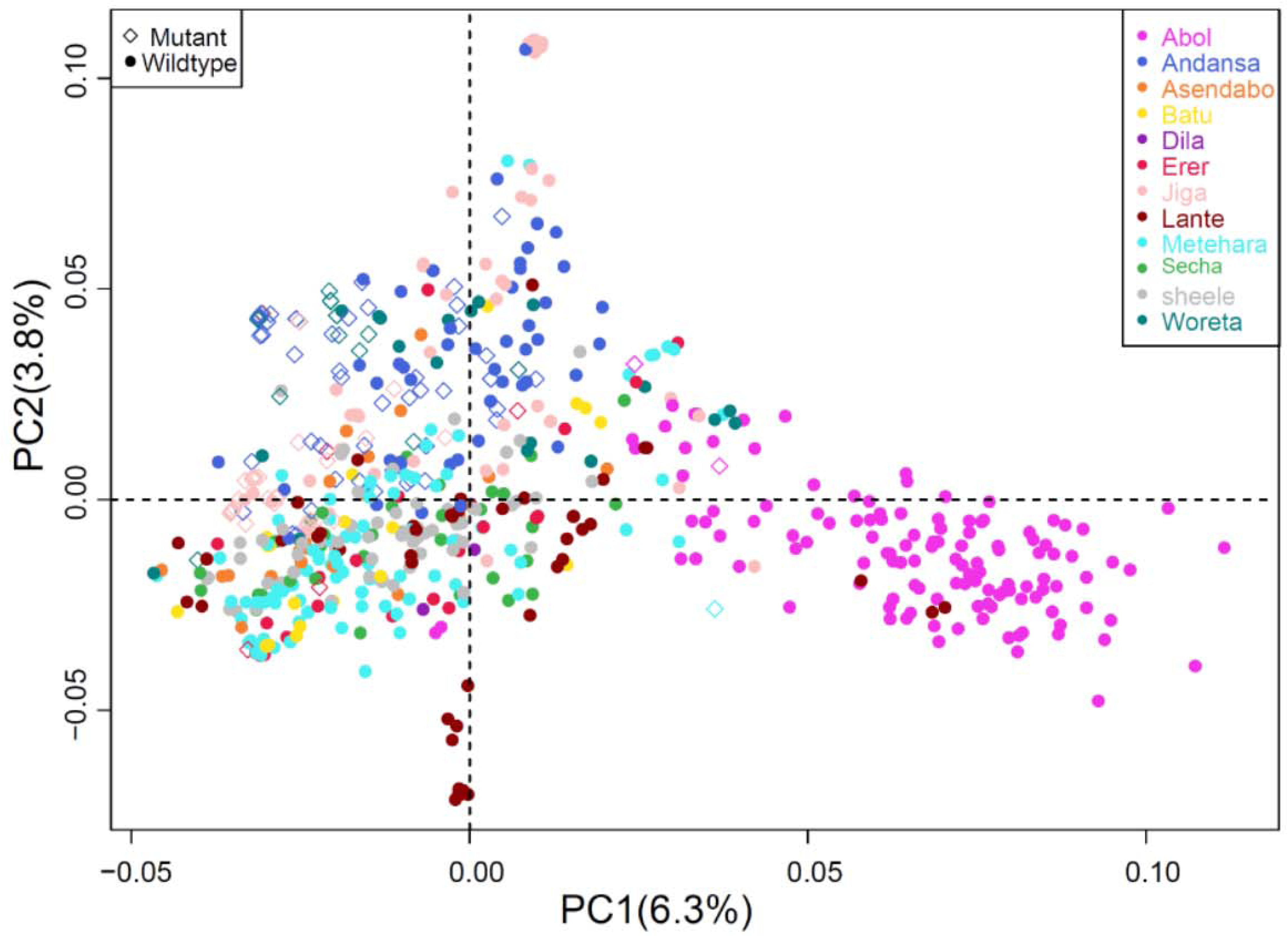
Population structure of *P. falciparum* populations per 622I mutation and Health facility level across five regions in Ethiopia. Colors indicate geographic sample origins and shapes indicate mutation status. Each diamond or dot indicates individual parasites. Percentage of variance explained by each PCA presented in each plot.

### Supplementary tables

**To assess and view easily all supplementary tables compiled in one excel sheet.**

**Table S1**. Details of samples sequenced per region, health facility and year of sample collection.

**Table S2.** Metadata and genotype file for all successfully sequenced samples for drug resistance loci. Only successfully sequenced samples were used for drug resistance marker prevalence estimates. 0=Reference call (Wildtype), 1=Alternative heterozygous call (Mixed mutant), 2=Alternative homozygous call (Pure mutant), −1=Missing call. Note that the number of samples successfully genotyped varies per locus drug resistance MIP panel.

**Table S3.** Prevalence of key mutation per health facility, n=number samples sequenced at each health center and % prevalence of key mutation.

**Table S4.** List of molecular inversion probes (IBC2CORE MIPs) designed targeting genome-wide geographic informative SNPs.

